# Characteristics of Walk-In Clinic Physicians and Patients in Ontario, Canada: A Cross-Sectional Study

**DOI:** 10.1101/2024.01.16.24301360

**Authors:** Lauren Lapointe-Shaw, Christine Salahub, Peter C. Austin, Li Bai, Sundeep Banwatt, Simon Berthelot, R. Sacha Bhatia, Cherryl Bird, Laura Desveaux, Tara Kiran, Aisha Lofters, Malcolm Maclure, Danielle Martin, Kerry A. McBrien, Rita K. McCracken, J. Michael Paterson, Bahram Rahman, Jennifer Shuldiner, Mina Tadrous, Braeden A. Terpou, Niels Thakkar, Ruoxi Wang, Noah M. Ivers

**Affiliations:** University Health Network, Toronto, Ontario, Canada; ICES, Toronto, Ontario, Canada; Institute of Health Policy, Management and Evaluation, University of Toronto, Toronto, Ontario, Canada; Department of Medicine, University of Toronto, Toronto, Ontario, Canada; Women’s College Institute for Health System Solutions and Virtual Care, Women’s College Hospital, Toronto, Ontario, Canada; Division of General Internal Medicine and Geriatrics, University Health Network and Sinai Health System, Toronto, Ontario, Canada; Sunnybrook Research Institute, Toronto, Ontario, Canada; CarePoint Health, Mississauga, Ontario, Canada; Département de médecine de famille et de médecine d’urgence, Faculté de médecine, Université Laval, Québec City, Québec, Canada; Department of Cardiology, University Health Network, Toronto, Ontario, Canada; Patient partner, Toronto, Ontario, Canada; Institute for Better Health, Trillium Health Partners, Mississauga, Ontario, Canada; Department of Family and Community Medicine, University of Toronto, Toronto, Ontario, Canada; MAP Centre for Urban Health Solutions, St. Michael’s Hospital, Toronto, Ontario, Canada; Department of Family and Community Medicine, St. Michael’s Hospital, Unity Health Toronto, Toronto, Ontario, Canada; Peter Gilgan Centre for Women’s Cancers, Women’s College Hospital, Toronto, Ontario, Canada; Department of Anesthesiology, Pharmacology and Therapeutics, Faculty of Medicine, University of British Columbia, Vancouver, British Columbia, Canada; Department of Family Medicine, Women’s College Hospital, Toronto, Ontario, Canada; Departments of Family Medicine and Community Health Sciences, Cumming School of Medicine, University of Calgary, Calgary, Alberta, Canada; Department of Family Practice, University of British Columbia, Vancouver, British Columbia, Canada; Department of Family Medicine, McMaster University, Hamilton, Ontario, Canada; Primary Health Care Branch, Ministry of Health, Ontario, Canada; Department of Health Research Methods, Evidence, and Impact, McMaster University, Ontario, Canada; Leslie Dan Faculty of Pharmacy, University of Toronto, Toronto, Ontario, Canada; College of Nurses of Ontario, Toronto, Ontario, Canada

## Abstract

**Objective:** We aimed to describe family physicians who primarily practice in a walk-in clinic setting and compare them to family physicians who provide longitudinal care.

**Design:** A cross-sectional study that linked results from an annual physician survey (2019) to administrative healthcare data from Ontario, Canada. We compared the characteristics, practice patterns, and patients of physicians primarily working in a walk-in clinic setting, with family physicians providing longitudinal care.

**Setting:** Ontario, Canada.

**Participants:** Physicians who primarily worked in a walk-in clinic setting in 2019, as indicated by an annual physician survey.

**Exposure:** Whether the physician was a walk-in clinic physician or a family physician who provided longitudinal care.

**Main Measures:** Physician demographic and practice characteristics, as well as their patients’ demographic and healthcare utilization characteristics.

**Results:** Compared to the 9,137 family physicians practicing longitudinal care, the 597 physicians who self-identified as practicing primarily in walk-in clinics were more frequently male (67% vs. 49%) and could speak a language other than English or French (43% vs. 32%). Walk-in clinic physicians had more encounters with patients who were younger (*M* 37 vs. 47 years), had lower levels of prior healthcare utilization (15% vs. 19% in highest band), who resided in large urban areas (87% vs. 77%), and in highly ethnically diverse neighborhoods (45% vs. 35%). Walk-in clinic physicians had more encounters with unattached patients (32% vs. 17%) and with patients attached to another physician outside their group (54% vs. 18%).

**Conclusion:** Physicians who primarily work in walk-in clinics saw many patients from historically underserved groups, and many patients who were attached to another family physician.

## KEY POINTS

- This is the first study to compare walk-in clinic physicians to longitudinal practice physicians using a population-based survey.
- Walk-in physicians saw younger, healthier patients, who lived in large urban and ethnically diverse areas.
- verall, 33% of walk-in physicians’ encounters were to patients who did not have a regular family physician, and another 54% were already attached to another family physician.
- We estimated that the time walk-in clinic physicians spent working in walk-in clinics is similar to that which would be needed to attach a quarter of all unattached patients in Ontario.

## INTRODUCTION

One in 10 Canadians report that they do not have a regular family or other primary care physician,^1^ and many who do still struggle to get timely access.^2,3^ Walk-in clinics provide care to patients with and without a regular physician, and do not require a scheduled appointment or an ongoing relationship between patient and provider.^4–6^ An estimated one-third of the population visited a walk-in clinic annually prior to the COVID-19 pandemic.^7^ Yet, walk-in clinics have been criticized for poor continuity of both relationships and information, which is associated with worse patient outcomes.^8,9^

Fewer family physicians are choosing to provide longitudinal primary care.^10–13^ In 2021, 1 in 5 family physicians in Toronto were considering closing their practices in the next 5 years, and only 5% indicated they were actively seeking to grow their practices.^14^ Reasons for leaving practice included health concerns, financial pressures, burnout, retirement, and other work options in or outside of medicine.^15–17^ As a result, access to longitudinal primary care is likely to worsen in the coming decade.^18^

For family physicians, walk-in clinics may offer a practice alternative. Yet, little is known about the characteristics of physicians who work primarily in a walk-in setting. Our primary study objective was to describe their characteristics, practice patterns and patient populations, and compare these to longitudinal family physicians. Given current crises in primary care access and health human resources, an additional objective was to translate walk-in clinic days worked to the number of patients that could become attached if this time were re-allocated to supporting longitudinal primary care.

## METHODS

### Study Design & Setting

This was a cross-sectional study of Ontario family physicians. Ontario is Canada’s most populous province, and in 2019 had 14.5 million residents and approximately 14,000 practicing family physicians.^19^ About 13% of the population (2 million people) does not have a regular family doctor.^20,21^

Primary care reforms over the past 15 years attached 81% of Ontarians to a family physician through a patient enrolment model.^22^ These models include an access-related bonus that is reduced by the total amount of fee-for-service charges made by enrolled patients attending outside clinics (e.g. walk-in clinics).^23–25^

### Data Sources

Health administrative datasets were linked using unique encoded identifiers and analyzed at ICES (**Appendix Table 1** lists databases used). ICES is an independent, non-profit research institute whose legal status under Ontario’s health information privacy law allows it to collect and analyze healthcare and demographic data, without consent, for health system evaluation and improvement.

Administrative healthcare data were linked (through a data sharing agreement) to the results of the 2019 College of Physicians and Surgeons of Ontario (CPSO) annual license renewal survey, which is a mandatory component of physician license renewal in Ontario.^26^ A mandatory question asks physicians to list all their practice settings and the hours worked in each setting, per week. The CPSO then derived the following variables: whether a “walk-in clinic or episodic care clinic outside of a hospital” was a setting where a physician worked: i) more than 0 hours per week, ii) the greatest number of hours per week (of all listed settings) and iii) the majority of their working time (50% or more of the member’s practice hours).

### Physician Population

We included all 2019 CPSO survey respondents with a specialty of family medicine or with no specialty reported, excluding those who could not be linked, not actively practicing in 2019, or with a practice pattern consistent with a focused or specialist practice, or who worked fewer than 44 days per year.^27^

### Physician Comparison Groups

We defined “walk-in clinic physicians” as those who spent the greatest number of hours worked and the majority of their time in this setting, and who had at least one day with 10 or more office encounters with patients who were not enrolled to them personally.

The comparison group was longitudinal family physicians who were not included in the above definition and who practiced comprehensive primary care, as defined by a standard ICES algorithm (**Appendix Table 2**).^27^

### Physician Characteristics

Physician and practice characteristics including years in practice, gender, and patient enrolment model type were included (all variables listed in **Appendix Table 2**). Physician-level continuity was defined as the proportion of all their enrolled or virtually rostered patients with whom they had encounters in 2019. Virtual rostering assigns patients who are not formally enrolled to the family physician with the most claims for primary care services in the previous year (**Appendix Table 2**).^22^

### Patient Characteristics at Encounters

We examined patient characteristics at each office encounter, from January to December 2019, excluding walk-in clinic physician encounters with their own enrolled patients. This included (all variables in **Appendix Table 2**) whether they were new insurance registrants in the past 10 years (a proxy for recent immigration to the province), level of material deprivation, dependency, instability, and ethnic concentration in patients’ residential neighbourhoods, divided into quintiles^28,29^, as well as count of comorbidities and previous healthcare utilization based on Aggregated Diagnosis Groups (ADGs) and Resource Utilization Bands (RUBs), both derived from The John Hopkins ACG® System Version 10.^30^ We also described the physician-patient relationship at each encounter, and the most frequent diagnoses.

### Analysis

We first described physicians and their encounters and plotted their main practice locations on a map of Ontario. As suggested by our patient partners, we overlaid a map of the density of unenrolled patients in 2019.

We compared the characteristics of walk-in and longitudinal FPs, and their encounters, using standardized mean differences (SMDs), with differences greater than 10% (0.1) considered meaningful.^31^

We estimated how many more Ontarians could be enrolled to a family physician if each walk-in clinic work day were reallocated to support a primary care enrolment model (methods in **Appendix Table 3**). The purpose of this was neither to simulate nor inform any particular intervention, but rather to appreciate the size of the walk-in clinic physician workforce, in the context of an Ontario population with over 2 million unattached patients.^20^ Analysis was executed using SAS software, version 9.4 (SAS Institute Inc., Cary, NC). Figures were generated using Tableau version 2022.3.7. We followed the STROBE reporting guidelines.^32^

### Ethics Approval

This study was approved by the Women’s College Hospital Research Ethics Board (REB 2020-0095-E) with a waiver of patient consent.

### Patient Partner Participation

Three patient partners who have been longitudinally involved in the project team’s work on walk-in clinics^33^ contributed to the analytic plan and results interpretation.

## RESULTS

In 2019, 16,337 family physicians completed the survey. After exclusions (**Figure 1**), 597 (5.7%) of the 10,443 included family physicians were classified as walk-in clinic physicians. Another 9,137 (87.5%) were longitudinal FPs, of whom 1,085 (11.9%) reported spending some time in a walk-in setting. Physicians who primarily worked in walk-in clinics had lower physician-level continuity than longitudinal FPs (0.1 vs. 0.6, SMD 1.98).

**Figure 1.**
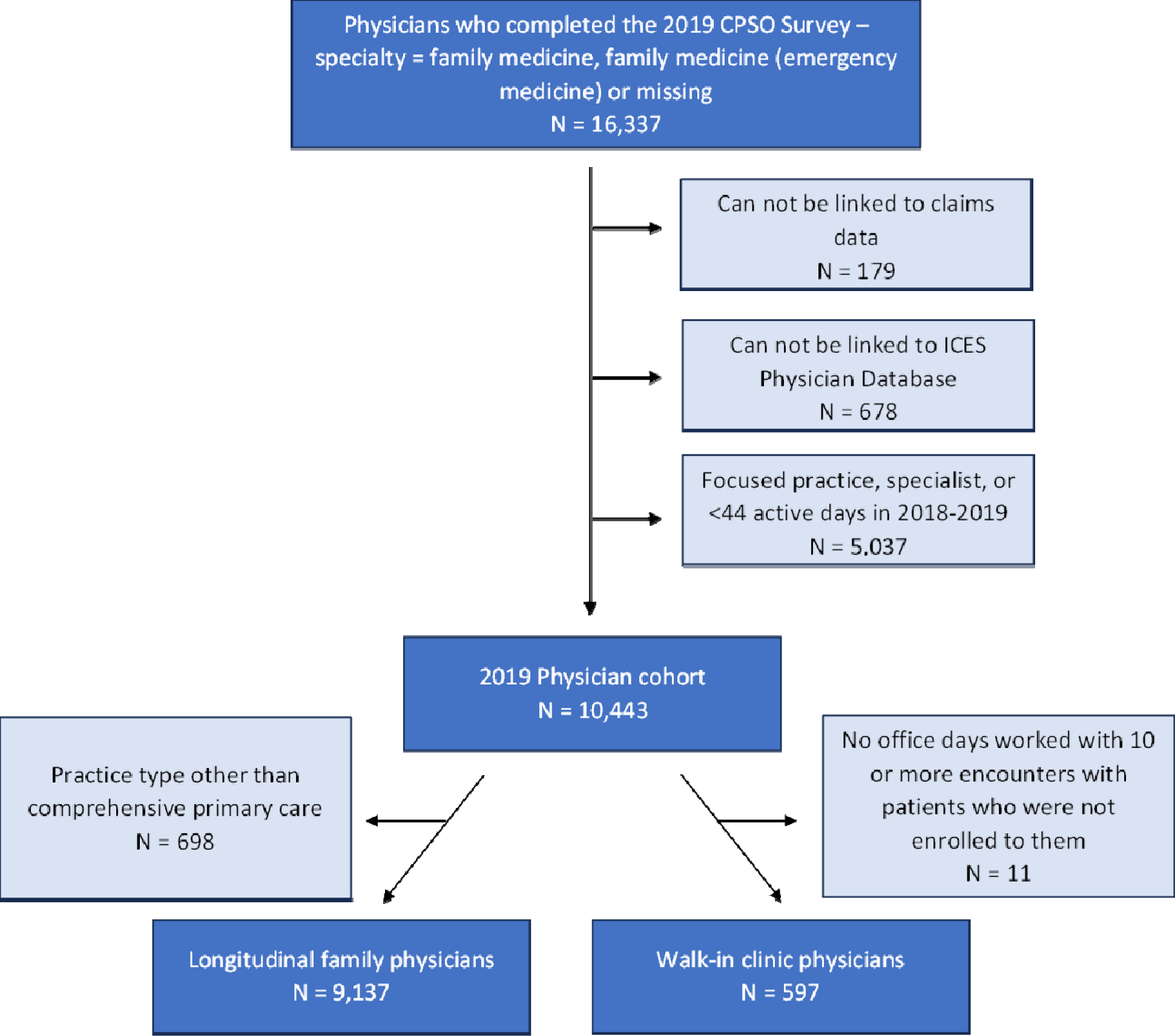
Flowchart of Family Physicians Included**. NB:** Physicians who met the definition for comprehensive practice and also practiced in a walk-in clinic setting some of the time, but did not meet the definition for “walk-in clinic physicians” listed above were included in the longitudinal family physician group.

### Physician Characteristics

Compared to longitudinal FPs, more walk-in clinic physicians were male (**Table 1**), offered services in a language other than English or French, and were more likely to practice in large urban areas. Walk-in physicians’ primary practice addresses were clustered in major urban areas, only covering some of the areas with the highest proportion of unattached patients (**Figure 2**).

**Table 1.** Characteristics of Walk-In Clinic vs. Longitudinal Family Physicians in 2019.

| Physician Characteristics | Walk-In Clinic Physicians<br>N = 597 | Longitudinal Family Physicians<br>N = 9,137 | Standardized Mean Difference |
| --- | --- | --- | --- |
| **Walk-in clinic setting is a practice setting, n (%)** | 597 (100) | 1,085 (11.9) | - |
| **Years in practice, n (%)** |  |  |  |
| Mean ± SD | 25.5 ± 14.8 | 23.8 ± 13.4 | 0.12 |
| Median (IQR) | 25 (13-37) | 24 (11-34) | NA |
| 0 to 5 | 51 (8.5) | 743 (8.1) | 0.02 |
| 6 to 10 | 76 (12.7) | 1,399 (15.3) | 0.07 |
| 11 to 20 | 118 (19.8) | 1,740 (19.0) | 0.02 |
| 21 to 30 | 131 (21.9) | 2,143 (23.5) | 0.04 |
| 31 plus | 221 (37.0) | 3,112 (34.1) | 0.06 |
| **Male, n (%)** | 399 (66.8) | 4,487 (49.1) | 0.37 |
| **Language Spoken, n (%)** |  |  |  |
| English and French | 47 (7.9) | 1,015 (11.1) | 0.11 |
| English and another language (not French) | 255 (42.7) | 2,888 (31.6) | 0.23 |
| English only | 295 (49.4) | 5,234 (57.3) | 0.16 |
| **Number of practice addresses, n (%)** |  |  |  |
| Mean ± SD | 1.8 ± 1.6 | 1.3 ± 0.7 | 0.36 |
| Median (IQR) | 1 (1-2) | 1 (1-1) | NA |
| 1 | 334 (55.9) | 7,115 (77.9) | 0.48 |
| 2 | 164 (27.5) | 1,474 (16.1) | 0.28 |
| 3 | 61 (10.2) | 382 (4.2) | 0.24 |
| 4+ | 38 (6.4) | 166 (1.8) | 0.23 |
| **Primary practice location, n (%)** |  |  |  |
| Large Urban | 543 (91.0) | 7,068 (77.4) | 0.38 |
| Small Urban | 16 (2.7) | 425 (4.7) | 0.11 |
| Rural | 34 (5.7) | 1,573 (17.2) | 0.37 |
| Missing | 4 (0.7) | 71 (0.8) | 0.01 |
| **Patient enrolment model type, n (%)** |  |  |  |
| Enhanced fee-for-service | 201 (33.7) | 2,538 (27.8) | 0.13 |
| Non-team capitation | 16 (2.7) | 2,742 (30.0) | 0.80 |
| Team capitation | 12 (2.0) | 2,479 (27.1) | 0.76 |
| Other Group | 0 | 85 (0.9) | 0.14 |
| No Group | 366 (61.3) | 1,250 (13.7) | 1.13 |
| Missing | 2 (0.3) | 43 (0.5) | 0.02 |
| **Percent of income from fee-for-service, n (%)** |  |  |  |
| Mean ± SD | 89.2 ± 20.5 | 44.1 ± 35.7 | 1.55 |
| Median (IQR) | 98 (89-100) | 26 (12-82) | NA |
| 0 to 25% | 22 (3.7) | 4,501 (49.3) | 1.21 |
| >25% to 50% | 21 (3.5) | 891 (9.8) | 0.25 |
| >50% to 75% | 31 (5.2) | 604 (6.6) | 0.06 |
| >75% to 100% | 521 (87.3) | 3,103 (34.0) | 1.30 |
| Missing | 2 (0.3) | 38 (0.4) | 0.01 |
| **Days worked in all locations, n (%)** |  |  |  |
| Mean ± SD | 201.9 ± 74.4 | 230.9 ± 58.0 | 0.44 |
| Median (IQR) | 210 (144-260) | 240 (200-271) | NA |
| Less than or equal to 60 days | 15 (2.5) | 88 (1.0) | 0.12 |
| 61 to 120 days | 90 (15.1) | 487 (5.3) | 0.33 |
| 121 to 180 days | 120 (20.1) | 1,039 (11.4) | 0.24 |
| 181 to 240 days | 166 (27.8) | 2,981 (32.6) | 0.11 |
| 241 to 300 days | 159 (26.6) | 3,843 (42.1) | 0.33 |
| 301 plus days | 47 (7.9) | 699 (7.7) | 0.01 |
| **Days worked in an office setting, n (%)** |  |  |  |
| Mean ± SD | 188.2 ± 75.6 | 184.8 ± 56.3 | 0.05 |
| Median (IQR) | 196 (125-250) | 190 (152-223) | NA |
| Less than or equal to 60 days | 33 (5.5) | 291 (3.2) | 0.12 |
| 61 to 120 days | 108 (18.1) | 979 (10.7) | 0.21 |
| 121 to 180 days | 123 (20.6) | 2,674 (29.3) | 0.20 |
| 181 to 240 days | 160 (26.8) | 3,978 (43.5) | 0.36 |
| 241 to 300 days | 141 (23.6) | 1,123 (12.3) | 0.30 |
| 301 plus days | 32 (5.4) | 92 (1.0) | 0.25 |
| **Number of patients seen in office in 2019** |  |  |  |
| Mean ± SD | 4,112 ± 3,091 | 1,521 ± 1,152 | 1.11 |
| Median (IQR) | 3,475 (1,839-5,537) | 1,218 (855-1,750) | NA |
| **Number of office encounters in 2019** |  |  |  |
| Mean ± SD | 6,514 ± 5,130 | 3,788 ± 2,839 | 0.66 |
| Median (IQR) | 5,379 (2,780-8,884) | 3,024 (1,985-4,687) | NA |
| **Median number of patients seen in a day of office encounters** |  |  |  |
| Mean ± SD | 31.6 ± 17.1 | 19.2 ± 10.6 | 0.88 |
| Median (IQR) | 29 (20-40) | 17 (12-23) | NA |
| 0 to 15, n (%) | 78 (13.1) | 3,977 (43.5) | 0.72 |
| 16 to 30, n (%) | 249 (41.7) | 4,014 (43.9) | 0.05 |
| 31 to 45, n (%) | 169 (28.3) | 832 (9.1) | 0.51 |
| 46 to 60, n (%) | 65 (10.9) | 205 (2.2) | 0.35 |
| 61 plus, n (%) | 33 (5.5) | 69 (0.8) | 0.28 |
| Missing, n (%) | 3 (0.5) | 40 (0.4) | 0.01 |
| **Physician-level continuity, n (%)** |  |  |  |
| Mean ± SD | 0.1 ± 0.2 | 0.6 ± 0.3 | 1.98 |
| Median (IQR) | 0 (0-0) | 1 (0-1) | NA |
| 0 to 0.1 (Lowest) | 455 (76.2) | 1,455 (15.9) | 1.52 |
| >0.1 to 0.3 | 94 (15.7) | 625 (6.8) | 0.28 |
| >0.3 to 0.5 | 32 (5.4) | 1,031 (11.3) | 0.22 |
| >0.5 to 0.7 | 8 (1.3) | 1,796 (19.7) | 0.63 |
| >0.7 to 0.9 | 8 (1.3) | 3,132 (34.3) | 0.95 |
| >0.9 (Highest) | 0 | 1,032 (11.3) | 0.51 |
| No visits | 0 | 66 (0.7) | 0.12 |
| **Physician and group-level continuity, n (%)** |  |  |  |
| Mean ± SD | 0.1 ± 0.2 | 0.7 ± 7.6 | 0.12 |
| Median (IQR) | 0 (0-0) | 1 (0-1) | NA |
| 0 to 0.1 (Lowest) | 454 (76.0) | 1,440 (15.8) | 1.52 |
| >0.1 to 0.3 | 91 (15.2) | 591 (6.5) | 0.29 |
| >0.3 to 0.5 | 33 (5.5) | 906 (9.9) | 0.17 |
| >0.5 to 0.7 | 9 (1.5) | 1,675 (18.3) | 0.59 |
| >0.7 to 0.9 | *5-9 | 3,119 (34.1) | 0.96 |
| >0.9 (Highest) | *1-5 | 1,340 (14.7) | 0.56 |
| No visits | 0 | 66 (0.7) | 0.12 |

**Figure 2.**
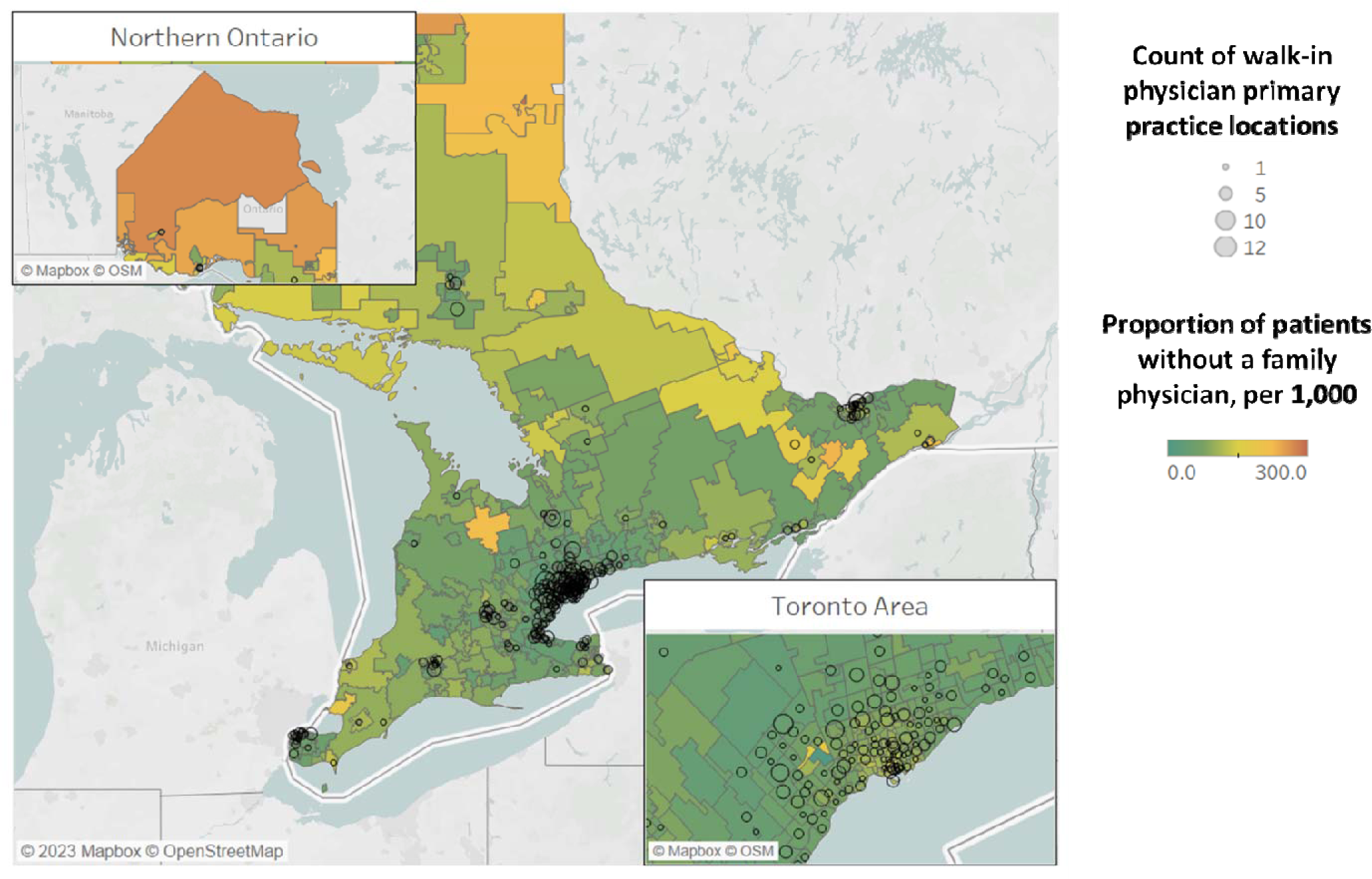
Proportion of Patients Without a Family Physician (per 1,000 Patients) Overlaid with Walk-In Clinic Physician Primary Practice Locations in Ontario, Canada. *Note.* Circle size reflects the number of walk-in clinic physicians with a primary practice location within a forward sortation area (FSA, first 3 digits of postal code). Colour of FSA reflects the proportion of patients without a family physician (non-enrolled) per 1,000 patients on a scale from 0 to 300. n = 11 physicians were missing an FSA and n = 2 physicians had a primary practice location outside of Ontario (not plotted). Total N = 584 physicians.

Of the walk-in clinic physicians who had enrolled patients (38.5%), most worked in enhanced fee-for-service models (87.8%). Walk-in clinic physicians received a mean of 89.2% of their income from fee-for-service billings, compared to 44.1% for longitudinal FPs (SMD 1.55).

Compared to longitudinal FPs, walk-in clinic physicians worked a similar number of days in an office setting, but had 1.7 times as many office encounters, and saw almost three times as many patients.

### Encounter-level Characteristics

Walk-in clinic physicians provided 13% of all their office encounters to patients enrolled to them or their group (longitudinal = 64.7%, SMD 1.25), 33.0% of encounters to patients who were not formally enrolled to any family physician (longitudinal 17.0%, SMD 0.38), and 54.0% of encounters to patients who were enrolled to another physician outside their group (longitudinal 18.3%, SMD 0.80; **Table 2**).

**Table 2.**
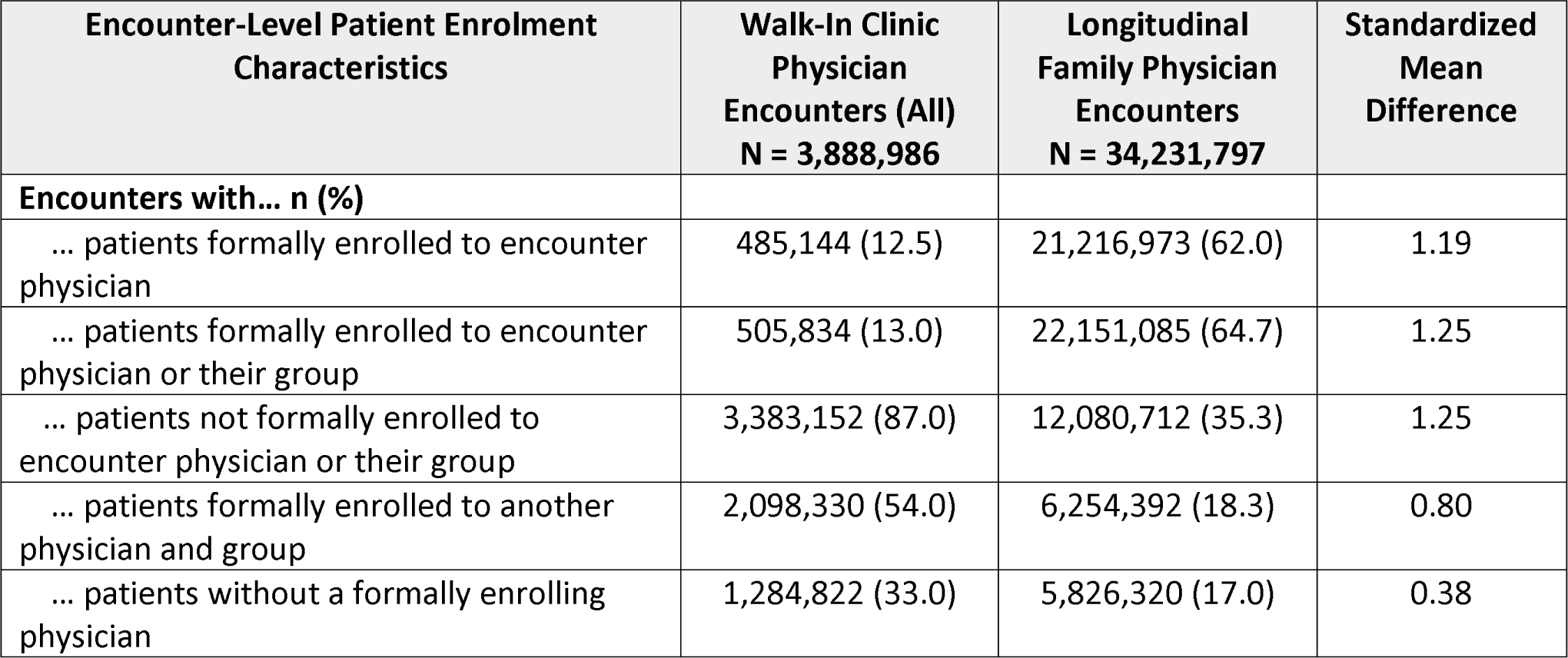
Encounter-Level Patient Enrolment Characteristics of Walk-In Clinic vs. Longitudinal Family Physicians in 2019.

Acute conditions such as the common cold, acute bronchitis, and acute sinusitis were more commonly diagnosed at encounters with walk-in clinic physicians; chronic conditions such as hypertension and diabetes were more commonly listed diagnoses for encounters with longitudinal FPs (**Table 3**).

**Table 3.** Top 10 OHIP Diagnoses Billed by Walk-In Clinic Physicians (walk-in encounters*), versus Longitudinal Family Physicians (all office encounters) in 2019.

| **Diagnoses from Walk-In Clinic Physician Encounters, n (%)**<br>**N = 3,427,511** | **Diagnoses from Longitudinal Family Physician Encounters**<br>**N = 34,231,797** |
| --- | --- |
| Acute nasopharyngitis, common cold<br>413,757 (12.1) | Essential, benign hypertension<br>2,175,602 (6.4) |
| Other ill-defined conditions<br>202,667 (5.9) | Mental health <sup>c</sup><br>1,960,003 (5.7) |
| Acute bronchitis<br>114,844 (3.4) | Diabetes mellitus including complications<br>1,864,273 (5.5) |
| Musculoskeletal symptoms other than back pain <sup>a</sup><br>101,327 (3.0) | Acute nasopharyngitis, common cold<br>1,757,705 (5.1) |
| Gastrointestinal symptoms <sup>b</sup><br>98,834 (2.9) | Other ill-defined conditions<br>1,561,174 (4.6) |
| Mental health <sup>c</sup><br>94,239 (2.8) | Musculoskeletal symptoms other than back pain <sup>a</sup><br>1,183,028 (3.5) |
| Essential, benign hypertension<br>89,947 (2.6) | Gastrointestinal symptoms <sup>b</sup><br>999,210 (2.9) |
| Acute sinusitis<br>88,064 (2.6) | Eczema, dermatitis<br>619,369 (1.8) |
| Eczema, dermatitis<br>78,698 (2.3) | Immunization – All types<br>589,202 (1.7) |
| Immunization – All types<br>76,312 (2.2) | Well baby care<br>587,608 (1.7) |

\* Excludes encounters with their enrolled patients
<sup>a</sup> Musculoskeletal symptoms other than back pain = “Leg cramps, leg pain, muscle pain, joint pain, arthralgia, joint swelling, masses”
<sup>b</sup> Gastrointestinal symptoms = “Anorexia, nausea and vomiting, heartburn, dysphagia, hiccough, hematemesis, jaundice, ascites, abdominal pain, melena, masses”
<sup>c</sup> Mental health = “Anxiety, neurosis, hysteria, neurasthenia, obsessive compulsive neurosis, reactive depression”

Walk-in encounter patients were, on average, 10 years younger than those of longitudinal FPs (**Table 4**). Walk-in clinic physicians had more encounters with patients who were recent registrants, from large urban areas, and from neighbourhoods with high ethnic diversity. Walk-in clinic physicians had fewer encounters with patients who had high previous healthcare utilization or who resided in neighbourhoods with high dependency scores.

**Table 4.** Encounter-Level Patient Characteristics of Walk-In Clinic Physicians (Non-Enrolled Patients Only) vs. Longitudinal Family Physicians (All Patients) in 2019.

| Encounter-Level Patient Characteristics | Walk-In Clinic Physician Encounters<br>N = 3,427,511 | Longitudinal Family Physician Encounters<br>N = 34,231,797 | Standardized Mean Difference |
| --- | --- | --- | --- |
| **Patient age in years at encounter** |  |  |  |
| Mean ± SD | 37.1 ± 21.2 | 47.2 ± 23.4 | 0.45 |
| Median (IQR) | 35 (22-53) | 50 (30-65) | NA |
| **Patient female sex, n (%)** | 1,922,428 (56.1) | 19,828,106 (57.9) | 0.04 |
| **Patient census-based neighbourhood income quintile, n (%)** |  |  |  |
| 1 (Lowest) | 766,860 (22.4) | 6,768,576 (19.8) | 0.06 |
| 2 | 701,452 (20.5) | 6,891,240 (20.1) | 0.01 |
| 3 | 717,575 (20.9) | 7,077,319 (20.7) | 0.01 |
| 4 | 661,496 (19.3) | 6,886,621 (20.1) | 0.02 |
| 5 (Highest) | 563,160 (16.4) | 6,471,876 (18.9) | 0.07 |
| Missing | 16,968 (0.5) | 136,165 (0.4) | 0.02 |
| **Patient dependency quintile, n (%)** |  |  |  |
| 1 (Lowest) | 1,238,611 (36.1) | 9,929,547 (29.0) | 0.15 |
| 2 | 754,396 (22.0) | 7,017,074 (20.5) | 0.04 |
| 3 | 547,554 (16.0) | 5,762,803 (16.8) | 0.02 |
| 4 | 464,452 (13.6) | 5,473,911 (16.0) | 0.07 |
| 5 (Highest) | 398,464 (11.6) | 5,807,265 (17.0) | 0.15 |
| Missing | 24,034 (0.7) | 241,197 (0.7) | 0 |
| **Patient neighbourhood material deprivation quintile, n (%)** |  |  |  |
| 1 (Lowest) | 689,057 (20.1) | 7,546,114 (22.0) | 0.05 |
| 2 | 687,760 (20.1) | 7,133,314 (20.8) | 0.02 |
| 3 | 647,660 (18.9) | 6,548,203 (19.1) | 0.01 |
| 4 | 661,218 (19.3) | 6,314,022 (18.4) | 0.02 |
| 5 (Highest) | 717,782 (20.9) | 6,448,947 (18.8) | 0.05 |
| Missing | 24,034 (0.7) | 241,197 (0.7) | 0 |
| **Patient neighbourhood ethnic diversity quintile, n (%)** |  |  |  |
| 1 (Lowest) | 205,441 (6.0) | 4,066,247 (11.9) | 0.21 |
| 2 | 334,319 (9.8) | 4,821,205 (14.1) | 0.13 |
| 3 | 514,920 (15.0) | 5,739,671 (16.8) | 0.05 |
| 4 | 818,525 (23.9) | 7,345,046 (21.5) | 0.06 |
| 5 (Highest) | 1,530,272 (44.6) | 12,018,431 (35.1) | 0.20 |
| Missing | 24,034 (0.7) | 241,197 (0.7) | 0 |
| **Patient neighbourhood residential instability quintile, n (%)** |  |  |  |
| 1 (Lowest) | 903,757 (26.4) | 8,224,984 (24.0) | 0.05 |
| 2 | 546,543 (15.9) | 6,191,466 (18.1) | 0.06 |
| 3 | 504,954 (14.7) | 5,907,334 (17.3) | 0.07 |
| 4 | 526,707 (15.4) | 5,813,679 (17.0) | 0.04 |
| 5 (Highest) | 921,516 (26.9) | 7,853,137 (22.9) | 0.09 |
| Missing | 24,034 (0.7) | 241,197 (0.7) | 0 |
| **Recent insurance registrant (<10 years), n (%)** | 430,960 (12.6) | 3,063,742 (8.9) | 0.12 |
| **Patient residence location, n (%)** |  |  |  |
| Large Urban | 2,989,435 (87.2) | 26,499,077 (77.4) | 0.26 |
| Small Urban | 35,625 (1.0) | 1,406,353 (4.1) | 0.20 |
| Rural | 287,350 (8.4) | 5,271,368 (15.4) | 0.22 |
| Missing | 103,253 (3.0) | 890,681 (2.6) | 0.03 |
| Unknown | 11,848 (0.3) | 164,318 (0.5) | 0.02 |
| **Patient resource utilization band, n (%)** |  |  |  |
| High | 495,295 (14.5) | 6,427,893 (18.8) | 0.12 |
| Moderate | 1,900,331 (55.4) | 20,049,104 (58.6) | 0.06 |
| Low | 1,031,885 (30.1) | 7,754,800 (22.7) | 0.17 |
| **Patient ADG score, n (%)** |  |  |  |
| High | 192,677 (5.6) | 1,973,973 (5.8%) | 0.01 |
| Moderate | 947,119 (27.6) | 9,837,567 (28.7%) | 0.03 |
| Low | 2,287,715 (66.7) | 22,420,257 (65.5%) | 0.03 |

### Walk-In Clinic Physician Workforce

We estimated that the walk-in clinic work hours provided by the 597 walk-in physicians included in our sample (median 3 days of walk-in work per physician-week, IQR 2-4, mean 3.1, SD 1.5), were equivalent to the physician-time needed to support the enrolment of a median 468,456 (IQR 314,247-712,308) additional patients to longitudinal primary care.

## INTERPRETATION

We found that 6% of family physicians licensed in Ontario in 2019 worked mainly in walk-in clinics. These physicians saw more patients daily and frequently treated unattached patients and those attached to an outside family physician. They were also more often male, multilingual, and practiced in multiple locations. Compared to longitudinal FPs, their patients were typically younger, less frequent healthcare users, from urban, ethnically diverse areas, or recent immigrants. This subset of family doctors worked an average of 3 days weekly in walk-in clinics, equivalent to the physician-time needed to enrol about 470,000 patients to a longitudinal FP.

Past studies of walk-in clinic physicians in Canada are more than a decade old. Previously, physicians working in walk-in clinics in Ontario and British Columbia were more often female.^6,34^ However, a study of 2015-2017 family medicine graduates found that more women preferred longitudinal primary care; this is potentially consistent with the finding of more men working in walk-in clinics in 2019.^35^

We found that, compared to longitudinal FPs, walk-in clinic physicians served more ethnically diverse populations. Recent immigrants have lower rates of attachment,^23^ cancer screening, ^36–38^ and care for chronic diseases.^39–43^ Whereas U.S. retail clinics typically serve higher income areas with lower levels of minority populations,^44–48^ walk-in clinics have acted as healthcare hubs for immigrant communities in Scandinavia.^49,50^ We found that a higher proportion of walk-in physicians offer services in a language other than English or French. They may be the children of immigrants, or immigrants themselves, in which case they could be in a better position to communicate with and understand the needs of newcomer patients.^51,52^ Nonetheless, newcomers should be offered the opportunity to also engage in longitudinal primary care to better meet their preventive and chronic care needs.

Timely access to episodic care remains foundational to a high-performing healthcare system. Ideally, this would be achieved in a setting that ensures continuity with longitudinal care. More than half of walk-in clinic physician encounters were with patients who were attached to an outside physician. In a survey study, 50% of patients indicated poor access to their regular family physician as the reason for visiting a walk-in clinic, although geographic proximity and general convenience were also commonly cited reasons.^53^ Although access challenges^2^ continue to drive demand for walk-in clinics in Canada, other countries such as Norway, the Netherlands, Sweden and Finland have few to no walk-in clinics.^54–57^ In these countries, there is greater accountability for same-day and after-hours access for urgent primary care issues, obviating the need for separate walk-in clinics.^58–63^

In translating total walk-in clinic workdays to the number of additional patients who could be enrolled, our aim was to contextualize the size of the walk-in physician workforce. The combined walk-in clinic work days reflect a workforce similar to that which would be needed to enrol a quarter of all unattached patients in Ontario. However, any potential intervention could more realistically aim for 10% or less of walk-in days reallocated, and should account for what is presently a mismatch between the geographic locations of walk-in physicians’ practices and many unattached patients.

In qualitative research from 20 years ago, family physicians reported walk-in clinics as necessary to fill the gap between primary care and emergency services.^64^ Several recent physician workforce trends have the potential to increase that gap, by decreasing access to longitudinal primary care. Fewer new family medicine graduates are choosing to pursue longitudinal family practice,^10,11^ and more are leaving practice.^14^ In an upcoming study of walk-in clinic physicians’ perspectives, scheduling flexibility, a desire to practice episodic care, the demand for walk-in care, and the fee-for-service payment model motivated the choice to practice in a walk-in setting.^65^ Paradoxically, physician workforce diversion to walk-in clinics and other alternatives may further reduce access to longitudinal primary care, ensuring that demand for walk-in clinics remains high.

Our study has several limitations. First, we relied on self-report to identify walk-in clinic physicians in Ontario. Physicians may underreport full-time walk-in clinic status, and as a result, our sample of walk-in clinic physicians is likely to be more specific than sensitive. Second, longitudinal physicians included some who reported working in a walk-in clinic setting part-time. We focused on those who primarily practiced in walk-in clinic settings to increase confidence that encounters with unenrolled patients were indeed walk-in clinic encounters. Walk-in style practice exists on a spectrum, and any misclassification of physicians in either direction would have biased our findings toward the null. Third, the physician primary practice address may not always correspond to a walk-in clinic location. Fourth, some communities and care teams were not included in our study as they do not appear in claims-based data. This includes Indigenous communities,^66^ as well as Community Health Centres, which care for under 2% of the population.^14^ Finally, changes in walk-in clinic physician services since 2019 could not be determined, but access to longitudinal, comprehensive primary care has worsened since the pandemic.^67,68^

## Conclusion

Physicians who primarily work in walk-in clinics fill gaps in the system by seeing unattached and underserved patients, albeit in an episodic model that is not designed to support preventive care, or the ongoing management of chronic disease. Over half of walk-in clinic physicians’ encounters are with patients who were enrolled to another practice, suggesting that access barriers could explain some of this walk-in clinic use.

## Data Availability

The dataset from this study is held securely in coded form at ICES. While legal data sharing agreements between ICES and data providers (e.g., healthcare organizations and government) prohibit ICES from making the dataset publicly available, access may be granted to those who meet pre-specified criteria for confidential access, available at www.ices.on.ca/DAS. The full dataset creation plan and underlying analytic code are available from the authors upon request, understanding that the computer programs may rely upon coding templates or macros that are unique to ICES and are therefore either inaccessible or may require modification.

