## Appendix for "Characteristics of Walk-In Clinic Physicians and Patients in Ontario, Canada: A Cross-Sectional Study"

**Table 1.** ICES data sources.

| **Database name** | **Description** |
| --- | --- |
| **College of Physicians and Surgeons of Ontario (CPSO) Annual Renewal Survey** | Each year, registered physicians in Ontario are required to submit their annual renewal online, which includes a completed renewal survey.^2^ This survey includes information on physician demographics, practice locations, and practice types. The 2019 CPSO survey information for this project was acquired through a data-sharing agreement between the CPSO and ICES. |
| **Discharge Abstract Database (DAD)** | Information on all admissions (excluding designated mental health beds) to acute care hospitals in Ontario. This includes dates of admission as well as diagnostic and procedural codes. Overall, diagnostic codes were found to be 82% sensitive for primary diagnosis when verified against chart abstraction.^3^ |
| **ICES Physician Database (IPDB)** | Demographic and practice information of physicians and surgeons licenced to practice in Ontario.^4^ |
| **National Ambulatory Care Reporting System (NACRS)** | Includes information for all emergency department visits since 2000. A re-abstraction study of diagnostic codes found 85% agreement for the main presenting problem.^5^ |
| **Ontario Health Insurance Plan (OHIP)** | Contains information on all billing claims submitted by Ontario physicians (consultations and procedures). Fee-for-service is the primary method of remuneration for 95% of specialist physicians and 50% of primary care physicians in Ontario. However, physicians practicing in non fee-for-service models submit shadow billings to OHIP, which appear as billing claims with a payment value of $0.^6^ |
| **Ontario Marginalization Index (ON-Marg)** | A geographic area-based index that combines a wide range of demographic indicators into four distinct dimensions of marginalization:^7,8^   - Households and dwellings (Residential instability) - Material resources (Material deprivation) - Age and labour force (Dependency) - Racialized and newcome populations (Ethnic concentration) |
| **Primary Care Population (PCPOP)** | Population-level dataset that includes all people in Ontario who are deemed alive and eligible to receive primary care at a given point in time- this relies on any healthcare encounter in Ontario in the previous 8 years. All indicators are as of the index date, with various lookback periods. The dataset contains information on demographics, primary care enrolment, and healthcare utilization over the previous 12 month period. The version of PCPOP used in the present study was from April 1^st^ 2019.^9^ |

**Table 2.** Operational definitions of all variables.

| **Variable** | **Data Source** | **Definition** |
| --- | --- | --- |
| Longitudinal family physicians | IPDB | Whether a physician primarily provided longitudinal, comprehensive primary care was defined by a standard ICES algorithm, described in Schultz and Glazier (2017).^10^ By this definition, longitudinal family physicians provided more than half their physician services in the area of outpatient primary care and these services fell into at least 7 of 22 activity areas, including general assessments, periodic health exams, mental health care, home visits, immunizations, Pap smears, and diabetes management. These physicians were by definition also not in the ‘walk-in clinic physicians’ group. |
| Years in practice | CPSO | Calculated as year of survey (2019) minus year of graduation from CPSO survey data. Categorized into the following groups:   - 0 to 5 years - 6 to 10 years - 11 to 20 years - 21 to 30 years - 30 plus years |
| Sex | CPSO | From CPSO, categorized as only: Female or Male |
| Language spoken | CPSO | Self-reported languages spoken categorized as:   - English only - English and French - English and another language |
| Number of practice addresses | IPDB | Count of practice addresses listed in the IPDB. |
| Primary practice location | IPDB | Postal code converted to Rurality Index of Ontario score^11^ of main practice address categorized as:   - Large urban (0 to 9) - Small urban (10 to 19) - Rural (20 plus) - Missing |
| Patient enrollment model type | PCPOP | Physician patient-enrollment model type categorized as below:   - Enhanced fee-for-service (family health group or comprehensive care model) - Non-team capitation (family health organization or family health network) - Team capitation (family health team) - Other group - Missing |
| Percent of income from fee-for-service | OHIP | Proportion of income that is fee-for-service, categorized as:   - 0 to 25% - >25% to 50% - >50% to 75% - >75% to 100% |
| Days worked in all locations | OHIP | Days worked per year with at least one encounter of any type in all locations for fiscal year 2018, for each physician. Categorized as:   - Less than or equal to 60 days - 61 to 120 days - 121 to 180 days - 181 to 240 days - 241 to 300 days - 301 plus days |
| Days worked in an office setting | OHIP | Same as above, except limited to office encounters, for each physician. |
| Number of patients seen in office in 2019 | OHIP | Number of patients seen in office by each physician in 2019. |
| Number of office encounters in 2019 | OHIP | Number of office encounters by each physician in 2019. |
| Median number of patients seen in a day of office encounters | OHIP | For each physician, the median count of patients seen per day, on days with at least one office visit. |
| Physician-level continuity | OHIP, PCPOP | ratio for each MD of:  - **Numerator:** all patients who were seen (office encounter) by this MD in 2018-2019 and who are either virtually or formally rostered to that MD in 2019  - **Denominator:** all unique patients who were seen by this physician (office encounters) in 2018 and 2019. |
| Physician and group-level continuity | OHIP, PCPOP | Same as above, however here:  -**Numerator:** all patients who were seen (office encounter) by this MD in 2018-2019 and who are either virtually or formally rostered to that MD’s group in 2019 |
| Patient age in years at encounter | OHIP | Age, in years, of patient at encounter. |
| Patient sex | OHIP | Sex at encounter categorized into male or female. |
| Patient census-based neighbourhood income quintile | PCPOP | Nearest census-based income quintile based on postal code (based on 2016 census, INCQUINT in PCPOP). |
| Patient dependency quintile | ON-Marg | A measure of the impacts of disability and dependence in an area.^8^ Includes:   - Proportion of the population who are aged 65 and older. - Dependency ratio (total population age 0 to 14 and 65+ divided by total population 15 to 64). - Proportion of the population not participating in labour force (aged 15+). |
| Patient material deprivation quintile | ON-Marg | An index of the inability for individuals and communities to access and attain basic material needs relating to housing, food, clothing, and education.^8^ Includes:   - Proportion of the population aged 25 to 64 without a high-school diploma. - Proportion of families who are lone parent families. - Proportion of total income from government transfer payments for population aged 15+. - Proportion of the population aged 15+ who are unemployed. - Proportion of the population considered low-income. - Proportion of households living in dwellings that are in need of major repair. |
| Patient neighbourhood ethnic diversity quintile | ON-Marg | A measure of the proportion of newcomers and/or non-white, non-Indigenous populations.^8^ Includes:   - Proportion of the population who are recent immigrants (past 5 years). - Proportion of the population who self-identify as a visible minority. |
| Patient residential instability quintile | ON-Marg | A measure of family and neighbourhood stability, including types and density of residential accommodations.^8^ Includes:   - Proportion of the population living alone. - Proportion of the population who are not youth (age 5-15). - Average number of persons per dwelling. - Proportion of dwellings that are apartment buildings. - Proportion of the population who are single/divorced/widowed. - Proportion of dwellings that are not owned. - Proportion of the population who moved during the past 5 years. |
| Patient is recent insurance registrant in Ontario (<10 years) | PCPOP | Recent registrant within the past 10 years, used as a proxy for immigration (IMMIG in PCPOP). Missing values are children less than 10 years of age. |
| Patient residence location | PCPOP | Postal code converted to RIO score (RIOG in PCPOP^11^):  0-9: Large urban  10-39: Small urban  40+: rural |
| Patient resource utilization band | DAD, NACRS, OHIP | Using Resource Utilization Bands (RUBs), per the Johns Hopkins ACG® System Version 7, in 2 years prior to the index date. Categorized as:  Low: 0-2  Moderate: 3  High: 4-5 |
| Patient ADG score | DAD, NACRS, OHIP | Count of ACG System Aggregated Diagnosis Groups (ADGs, per the Johns Hopkins ACG® System Version 7, in 2 years prior to the index date. Categorized as:  Low: 0-5  Moderate: 6-9  High: 10+ |

**Table 3.** Description of the calculation to estimate total patients that could be enrolled to longitudinal family physician care if walk-in clinic physicians’ walk-in days were re-allocated to supporting this.

For the purpose of this estimation, we made several assumptions. Based on an examination of the typical count of office visits per day made by walk-in clinic physicians (median 29, IQR 20-40), as well as clinician (NMI, DM, TK, AL) knowledge of typical family medicine volumes, we defined a walk-in clinic work day as any day with a minimum of 10 office encounters with non-enrolled patients. For each physician, we summed the number of walk-in work days in the year, and divided this by 52 weeks/year, then rounded to the nearest integer to obtain each physician’s typical count of walk-in clinic days worked per week (reported in results).

We then divided this number by 5 working days/week to obtain a conservative measure of full-time equivalents (FTEs) allocated to walk-in clinic days for each physician, ranging from 0 to 1.0 (those who had more than 1.0 FTE of walk-in clinic days were rounded down to 1.0).

Next, for each physician, we sampled an estimated 1.0 FTE patient panel size from a lognormal distribution with parameters derived from existing literature on typical practice size of family physicians in Ontario (median 1,290 patients, IQR of 800-1859).^12^ We then multiplied this number by each physician’s FTE value (between 0 and 1), to obtain the estimated potential enrolled patient panel size for each walk-in clinic physician.

We then summed the total patients that could be enrolled across all physicians. We repeated these steps 10,000 times to obtain the median (IQR) total additional patients who could be enrolled.
